# Exome-based Preconception Carrier Screening for the Mexican Jewish Community; Yields and Implications

**DOI:** 10.64898/2026.10.21.26366106

**Authors:** Rivka Birnbaum, Kamila Lee, Teresa Metta-Harari, Sylvia Harari-Arakindji, Shiri Gershon-Naamat, Israel Antman, Jacobo Borovoy-Gurvich, Elias Elías Charua-Levy, Paola Mendelsberg-Fishbein, Carol Perelman, Sarah Weissman, Dov Popper, Tamar Harel, Orly Elpeleg, Talia Wegman-Ostrosky, Hagar Mor-Shaked

## Abstract

**Introduction:** Preconception carrier screening (PCS) reduces monogenic disease incidence, exemplified by a 90% decline in Tay-Sachs among Ashkenazi populations. The Mexican Jewish Community (MJC), an endogamous population, may have increased prevalence of population-specific genetic disorders. The added value of exome sequencing (ES) over standard panels remains unclear.

**Objectives:** To identify pathogenic/likely pathogenic (P/LP) variants in autosomal recessive (AR), X-linked (XL), and mitochondrial genes, evaluate secondary findings (SF), and define population-specific allele frequencies in the MJC.

**Methods:** From 2020-2025, 787 couples (1,574 individuals) underwent ES following counseling and consent. Genetic data was analyzed including SNVs, CNVs, and mitochondrial DNA. Reporting was limited to P/LP variants and SF.

**Results:** Shared AR carrier status was identified in 49 couples (6.2%): 18.4% associated with severe, 16.3% moderate, and 65.3% with mild conditions. In 71.4% of couples, both partners carried the same variant, consistent with endogamy within the cohort. Maternal findings were identified in 10 women (1.27%). Clinically actionable SF were found in 2.85% of individuals. ES identified 12 at-risk couples (24.4%), undetected by standard panels. Individual genomic profiling revealed 920 rare P/LP variants, including 146 novel alleles, underscoring the distinct genetic architecture of the MJC.

**Conclusion:** ES-based PCS improves detection of at-risk couples beyond conventional screening and supports its use in endogamous populations.

## INTRODUCTION

Carrier screening (CS) identifies individuals or couples at risk of having offspring affected by clinically relevant monogenic conditions. Performed preconceptionally and paired with genetic counseling, it supports informed reproductive decision-making by enabling carrier couples to consider options such as preimplantation genetic testing, donor gametes, adoption, or prenatal diagnosis, thereby enhancing reproductive autonomy and reducing time-sensitive decisions during pregnancy (1). Once limited to include only autosomal recessive (AR) and X-linked (XL) conditions in specific ethnic groups, CS is now recommended universally to improve detection of at-risk couples and has been shown to reduce the incidence of these disorders (2).

The first structured Population-based Carrier Screening (PCS) program, targeting Tay-Sachs disease in Ashkenazi Jewish populations, reduced incidence by 90% across USA, Israel and Canada (3). Advances in sequencing technology have since enabled expansion to hundreds of conditions. The American College of Medical Genetics and Genomics (ACMG) recommends Tier 3 screening for the general population, covering 97 AR and 17 XL conditions (with a carrier frequency ≥1/200 (4,5), while the American College of Obstetricians and Gynecologists (ACOG) advises testing for cystic fibrosis, spinal muscular atrophy, and hemoglobinopathies, supplemented by ancestry-based or pan-ethnic screening for conditions with a frequency ≥1/100 (5). These distinct recommendations reflect the ongoing debate regarding the balance between comprehensiveness and clinical utility in genetic screening.

Next-generation sequencing (NGS) has transformed PCS by allowing rapid, cost-effective, and large-scale detection of both common and rare variants (2). Belnap et al. (2024) reported that exome sequencing (ES) identified 17 at-risk couples out of 150 (11.33% of the screened couples cohort), for AR or XL disorders, whereas only three would have been detected using ACMG Tier 3 criteria (6).

The probability of autosomal recessive (AR) disease in offspring increases with the degree of consanguinity. The Mexican Jewish community (MJC) was established in Mexico in the early 20th century by Ashkenazi, Syrian-Mizrahi, and Sephardi Jewish populations originating from Europe and the Middle East (7). Consanguineous unions, maintained through cultural practices, remain relatively common in subsets of Jewish populations (8).

Such practices increase the frequency of recurrent, shared AR pathogenic variants (9), resulting in population-specific differences in disease prevalence driven in part by genetic isolation (8). Accordingly, expanded carrier screening beyond ACMG Tier 3, and when appropriate Tier 4, may provide additional clinical value in these populations. Tier 4 screening is recommended in the setting of known or suspected consanguinity, or when personal or family history is suggestive. It includes genes with carrier frequencies below 1 in 200, encompassing Tier 3, and gene content may vary by laboratory (4,5,7).

However, current population databases have important limitations. GnomAD version 3.1 includes representation of Ashkenazi Jews but lacks Sephardi Jewish populations (10). Consequently, ancestry-specific carrier frequency estimates for Sephardi Jews derived from gnomAD may be limited or potentially misleading due to incomplete and non-specific population representation (11).

Morgenstern-Kaplan et al. (2017) used a 302-gene pan-ethnic panel in 82 MJC couples and identified five at-risk couples across three AR genes (8). A subsequent study by Dor Yeshorim in 2021, analyzed more than 3,800 individuals of Syrian Jewish ancestry and reported population-specific carrier frequencies for 40 AR variants, revealing both shared and unique alleles across Syrian, Iranian, and Ashkenazi Jewish groups (12). Despite these efforts, the added yield of exome-based carrier screening in these populations remains unknown.

ES represents a comprehensive approach for preconception carrier screening (PCS). It enables the simultaneous identification of pathogenic (P) and likely pathogenic (LP) variants in AR, XL, and mitochondrial genes, and provides the added advantage of detecting clinically actionable secondary findings (SF). These findings support the implementation of personalized preventive strategies, aligning with the principles of precision preventive medicine. Furthermore, it allows the characterization of population-specific variant frequencies within the MJC, which is essential for improving genetic counseling, understanding population structure, and informing community-based health initiatives. Therefore, the objectives of this study are to explore the genetic landscape of the MJC: (1) to detect couples at risk of having affected children by identifying P/LP variants in AR, XL, and mitochondrial genes; (2) to define population-specific allele frequencies, and (3) to assess clinically relevant SF.

## MATERIALS AND METHODS

### Participants

Couples were eligible to participate in the study if at least one member of the couple is of Jewish ethnic origin. No restrictions were applied regarding gender and age. All participants provided informed consent prior to enrollment in the protocol (CMABC-2361). Couples voluntarily participated with the objective of identifying shared recessive variants, as well as XL and mitochondrial variants, the last two limited to females. Ancestry was self-reported and categorized as Syrian, Sephardi, Ashkenazi, or mixed Jewish origin. All couples underwent pre-test genetic counseling, during which the benefits, risks, and limitations of exome sequencing were discussed. Results were returned in a post-test counseling session.

### Exome sequencing

Exome analysis was pursued on DNA extracted from the participants (whole blood). Exonic sequences from DNA were enriched with the SureSelect Human All Exon 50 Mb V5 Kit (Agilent Technologies, Santa Clara, CA, USA) or xGen Exome Research Panel IDT-V1 or IDT-V2 combined with xGen Human mtDNA Research Panel v1.0. Sequences were generated on a HiSeq2500 or NovaSeq6000 sequencing system (Illumina, San Diego, CA, USA) as 125-or 150-bp paired-end runs. The FASTQs were uploaded into the Geneyx Analysis platform (9). Alignment and variant calling of single nucleotide variations (SNVs), and copy number variations (CNVs) were called using Illumina DRAGEN Bio-IT. The resulting VCF files were comprehensively annotated on the Geneyx Analysis annotation engine, and presented for analysis, filtering and interpretation. The human genome assemblies hg19 (GRCh37) or hg38 were used as references. Exome analysis of the participants yielded 35-80 million reads, with a mean coverage of 55-100X.

### Shared carrier screening analysis

Combined exome analysis was performed for each couple, to assess shared carrier status. Only P/LP variants in AR/XL disease-causing genes were reported. Autosomal dominant (AD) variants were also evaluated and reported only in cases where variable expression or reduced penetrance has been described. Severity of the variants was determined based on the classification suggested by Lazarin et al.(10), into one of four groups: profound, severe, moderate, or mild. Mitochondrial analysis was also performed; reporting was limited to females. CNV analysis was performed for each couple, including the detection of compound heterozygous events involving both SNVs and CNVs.

### Secondary findings

Participants were given the option to receive information regarding SF or to decline disclosure, as documented through written informed consent. The reporting of SF followed the ACMG (SF v3.3, 2025) Recommendations for Reporting of Secondary Findings in Clinical Exome and Genome Sequencing, which currently encompasses 84 genes (11).

### Variant frequency

Variant frequencies were calculated for AR and XL genes harboring variants classified as P/LP. Only variants present in eight or more individuals within the cohort were included, corresponding to an estimated carrier frequency of 1 in 200, which represents the standard cutoff for carrier screening studies as recommended by the ACMG (Tier 3 Screening, Genetics in Medicine (7). The frequencies observed in the MJC were then compared with population frequencies reported in the gnomAD database.

### Exome diagnostic yield

To assess the diagnostic yield of exome analysis relative to existing population-based screening panels, we compared our results to the gene sets included in three established panels: Tier 3 ACMG-recommended guidelines, which includes 113 genes; the Dor Yeshorim published panel from 2021 (12). Dor Yeshorim is a community-based genetic screening initiative that anonymously evaluates carrier status to reduce the incidence of recessive disorders in Jewish populations; and the pan-ethnic panel previously applied to the MJC population by Morgenstern et al., which includes 302 AR genes (8).

### Database (DB) generation

A variant database was created by processing annotated VCF (Variant Call Format) files from subjects obtained through the Illumina DRAGEN pipeline. Annotated VCF files were converted into TSV (Tab-Separated Values) files to simplify data handling. The TSV files were filtered to retain only: (A) rare variants (>20 homozygous occurrences in gnomAD V2.1.1) that are published and considered P/LP variants on public databases such as ClinVar, or the Israeli National Genetic Database (INGD); or (B) variants with a Loss-of-function (LOF) effect, exhibiting no more than two homozygous and fewer than 700 heterozygous occurrences in gnomAD (V2.1.1). The frequency of each variant was then calculated across all individuals in the MJC population. Manual analysis was performed to ensure that all included variants are P/LP and in genes relevant to carrier screening. Finally, these variant counts were stored using Python’s Pandas DataFrame for efficient data manipulation. The code is available upon request.

## RESULTS

### Cohort Description

A total of 1,574 individuals (787 males and 787 females), participated in the study between 2020 and 2025 after providing informed consent. Of these, 54.25% (n=854) were self-reported as of Syrian origin, 11.24% (n=177) as Ashkenazi and 10.6% (n=167) as Syrian/Sephardi, while the remaining participants (23.88%; n = 376) reported mixed Jewish ancestry (Figure 1A).

**Figure 1.**
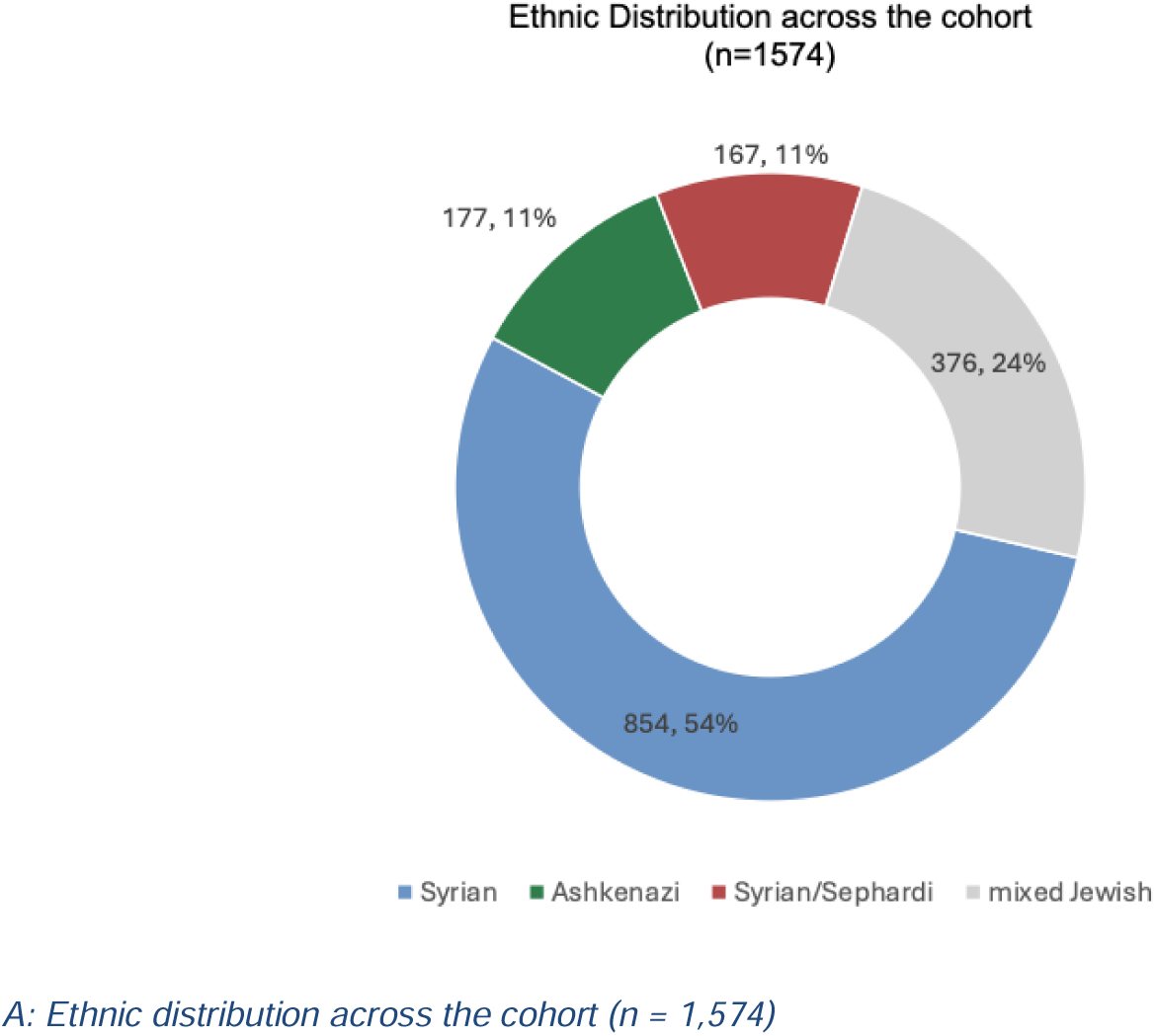
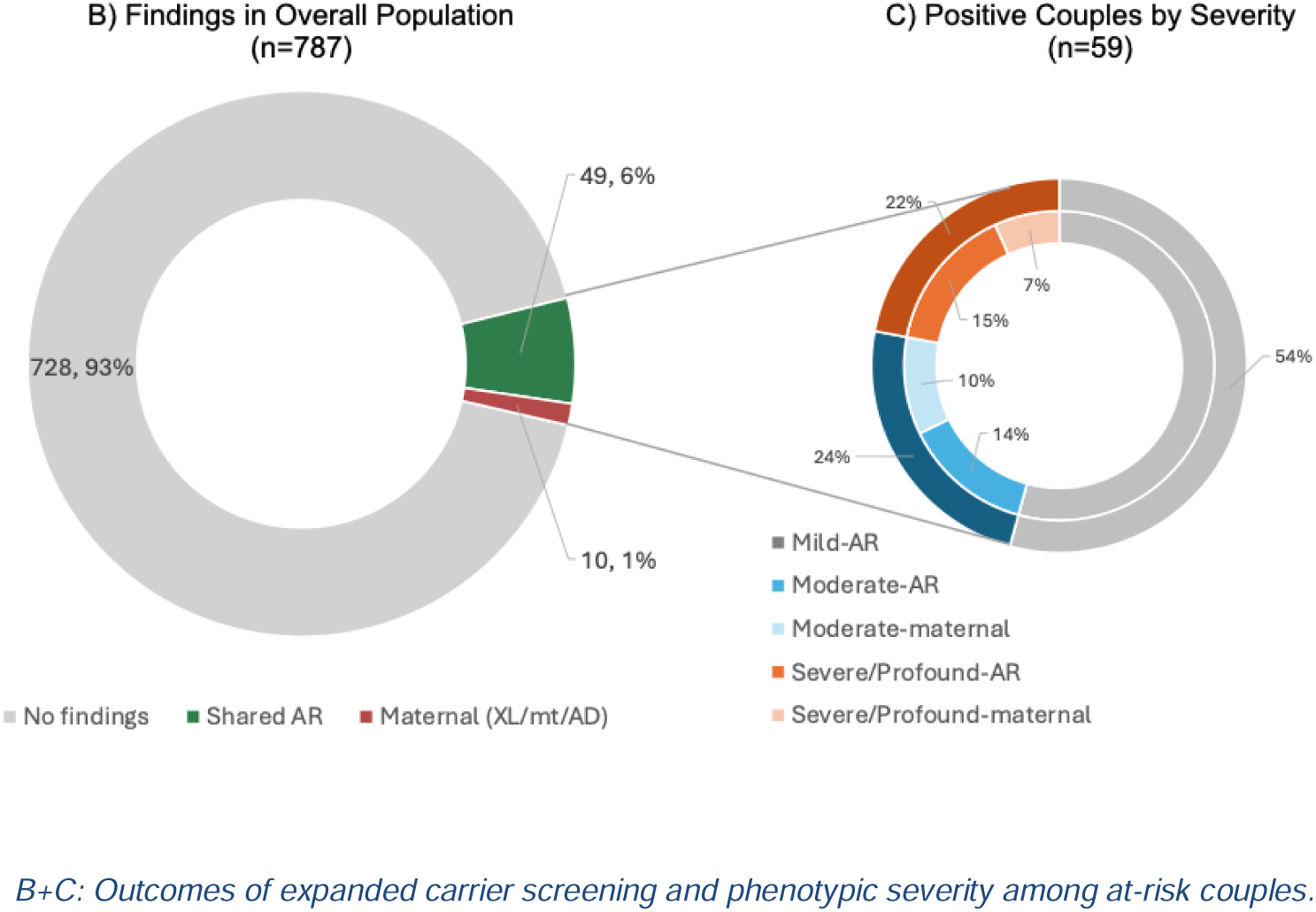
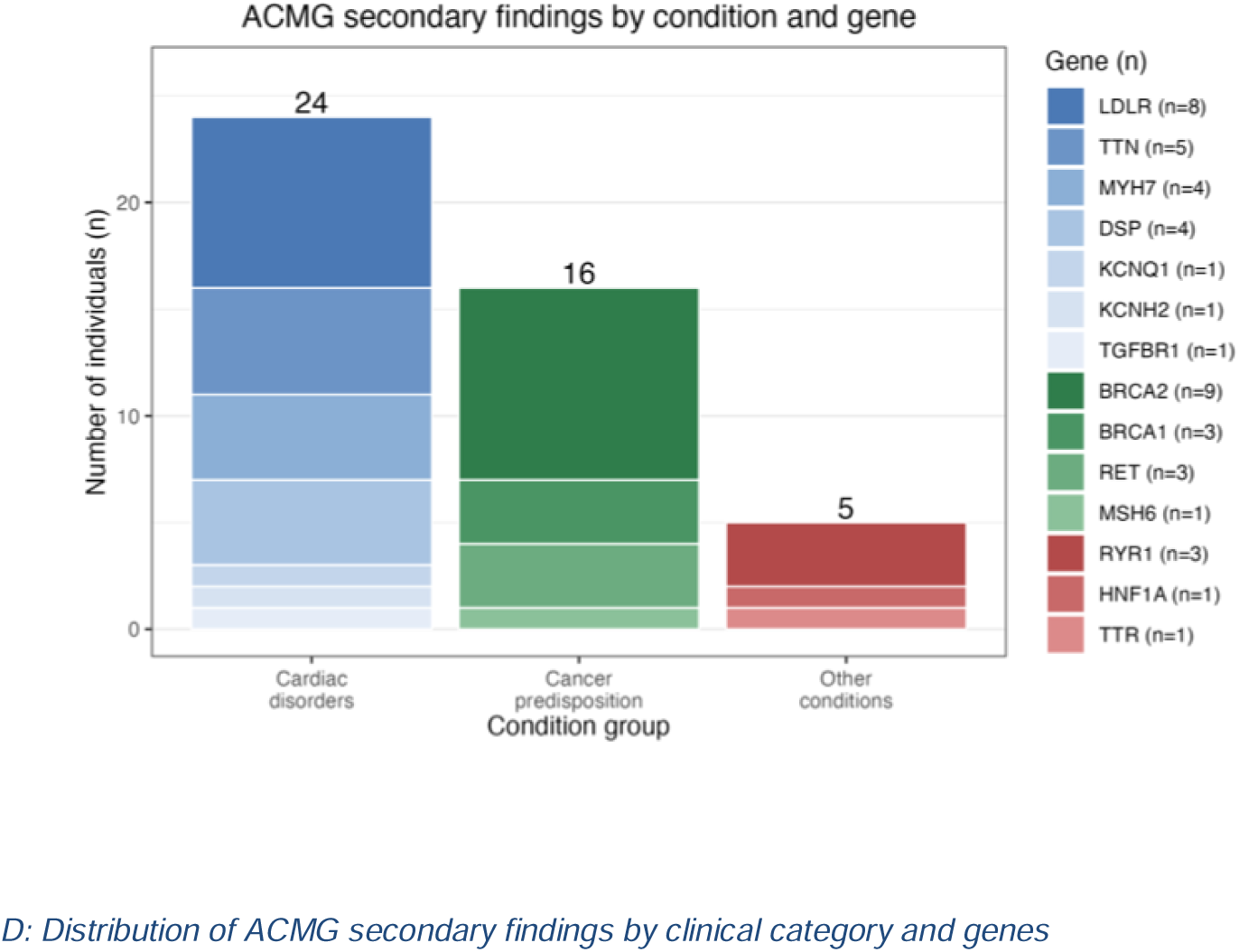
Ethnic Distribution, Carrier Screening Outcomes, and Secondary Findings in the Cohort. A) Donut plot depicting the distribution of self-reported ancestral backgrounds among all participants. The cohort is predominantly of Syrian Jewish ancestry (n = 854, 54%), with additional representation from mixed Jewish ancestry (n = 376, 24%), Syrian/Sephardi ancestry (n = 177, 11%), and Ashkenazi Jewish ancestry (n = 167, 11%). B) Donut plot summarizing results for all screened couples (n = 787). The vast majority had no shared pathogenic or likely pathogenic variants, whereas 49 couples (6%) were concordant carriers for the same autosomal recessive (AR) condition (“Shared AR carrier status”) and 10 couples (1%) had maternal pathogenic or likely pathogenic variants in X-linked, mitochondrial, or autosomal dominant (XL/mt/AD) genes. C) Donut plot illustrating clinical severity classification among all at risk couples (n = 59), categorized as mild (32 couples; AR), moderate (8 AR and 6 maternal XL/mt/AD), or severe/profound (9 AR and 4 maternal XL/mt/AD) based on established disease severity frameworks. D) Stacked bar chart showing the number of individuals with reportable secondary findings according to ACMG guidelines, stratified by major clinical category (cardiac disorders, cancer predisposition, and other conditions) and by gene. A total of 24 individuals harbored variants in genes associated with cardiac disorders (including *LDLR* [n = 8], *TTN* [n = 5], *MYH7* [n = 4], *DSP* [n = 4], *KCNQ1* [n = 1], *KCNH2* [n = 1], and *TGFBR1* [n = 1]), 16 individuals had variants in cancer predisposition genes (*BRCA2* [n = 9], *BRCA1* [n = 3], *RET* [n = 3], *MSH6* [n = 1]), and 5 individuals had variants in other actionable genes (*RYR1* [n = 3], *HNF1A* [n = 1], *TTR* [n = 1]).

The mean age was 27 years ± 3.80 (ranged between 18-44), and all individuals self-reported good general health.

### Shared Carrier Status

A total of 6.2% (n=49) of couples were identified as shared carriers of P/LP variants in AR disease-causing genes ranging from mild to profound clinical severity. These cases involved 17 distinct genes and 15 different disorders. Of these, 18.4% (n=9) were carriers of variants classified as associated with severe to profound conditions (Table 1, Figure 1B-C).

**Table 1.** Reported Shared AR variants. Summary of the 49 couples in whom both partners shared P/LP variants in AR inherited disorders, across 17 distinct genes and 15 associated disorders. The table includes gene symbol, associated disorders, variant information, predicted effect, and classification of both genetic variant and disease severity (mild, moderate, or severe/profound according to Lazarin et al.(10)). Abbreviation: ACMG -American College of Medical Genetics.

| Gene | Omim name | Omim # | Transcript | HGVSc | HGVSp | dbSNP | ACMG classification | # Affected couples | Effect | Severity Classification\ |
| --- | --- | --- | --- | --- | --- | --- | --- | --- | --- | --- |
| *DOCK7* | Developmental and epileptic encephalopathy 23 | 615859 | NM_001367561.1 | c.1832_1835del | p.(Val611AspfsTer32) | rs1571649028 | LP | 1 | Frameshift | profound |
| *ALDOB* | Hereditary fructose intolerance | 229600 | NM_000035.4 | c.448G>C | p.(Ala150Pro) | rs1800546 | PAT | 1 | Missense | severe |
| *ATP7B* | Wilson disease | 277900 | NM_000053.4 | c.845del | p.(Leu282ProfsTer2) | rs193922111 | PAT | 1 | Frameshift | severe |
| *COL6A2* | Ullrich congenital muscular dystrophy 1 | 254090 | NM_001849. | c.1402C>T | p.(Arg468Ter) | rs374669775 | PAT | 1 | Stop gain | severe |
| *PAH* | Phenylketonuria | 261600 | NM_000277.3 | c.1066-11G>A | N/R (intronic) | rs5030855 | PAT | 1 | Intronic | severe |
|  |  |  |  | c.898G>T | p.(Ala300Ser) | rs503page0853 | PAT |  | Missense |  |
| *PAH* | Phenylketonuria | 261600 | NM_000277.3 | c.688G>A | p.(Val230Ile) | rs62516152 | PAT | 1 | Missense | severe |
|  |  |  |  | c.727C>T | p.((Arg243*)) | rs5030846 | PAT |  | Stop gain |  |
| *PAH* | Phenylketonuria | 261600 | NM_000277.3 | c.898G>T | p.(Ala300Ser) | rs5030853 | PAT | 1 | Missense | severe |
|  |  |  |  | c.1208C>T | p.(Ala403Val) | rs5030857 | PAT |  | Missense |  |
| *PAH* | Phenylketonuria | 261600 | NM_000277.3 | c.829T>C | p.(Tyr277His) | rs78655458 | LP | 1 | Missense | severe |
|  |  |  |  | c.1243G>A | p.(Asp415Asn) | rs62644499 | PAT |  | Missense |  |
| *TGDS* | Catel-Manzke syndrome | 616145 | NM_014305.4 | c.281_282del | p.(Ile94SerfsTer19) | rs1889177568 | LP | 1 | Frameshift | severe |
| *CLCNKB + CLCNK1* | Bartter syndrome, type 3 , Bartter syndrome, type 4b, digenic | 613090, 607364 | NM_000085.5 | CLCNKA - del exons 4-20 , CLCNKB del exons 1-3 Del Chr1:16,351,257-16,372,181 | N/R | N/R | PAT | 1 | Deletion | moderate |
| *CLCNKB* |  |  |  | c.1830G>A | p.(Trp610*) | rs121909136 | PAT |  | Stop gain |  |

Table 1-continued
| Gene | Omim name | Omim # | Transcript | HGVSc | HGVSp | dbSNP | ACMG classification | # Affected couples | Effect | Severity Classification\ |
| --- | --- | --- | --- | --- | --- | --- | --- | --- | --- | --- |
| *FYCO1* | Cataract 18 | 610019 | NM_024513.4 | c.3150+1G>T | N/R (intronic) | rs766522434 | PAT | 1 | Splice site donor | moderate |
| *GBA* | Gaucher disease, perinatal lethal ; (types I ; II ; III ; IIIC) | 608013 ; 230800 ; 230900 ; 231000 ; 231005 | NM_000157.4 | c.1448T>C | p.(Leu483Pro) | rs421016 | PAT | 1 | Missense | moderate |
|  |  |  |  | c.1604G>A | p.(Arg535His) | rs75822236 | PAT |  | Missense |  |
| *GJB2* | Digenic deafness GJB2/GJB6 | 604418 | NM_004004.6 | c.101T>C | p.(Met34Thr) | rs35887622 | PAT | 1 | Missense | moderate |
| *GJB6* |  |  | NM_001110219 | Exons 1-5 deletion Del Chr13:20,796,833-21,086,689 | N/R | N/R | PAT |  | Deletion |  |
| *SEC23B* | Congenital dyserythropoietic anemia type II | 224100 | NM_006363.6 | c.325G>A | p.(Glu109Lys) | rs121918221 | PAT | 1 | Stop gain | moderate |
| *TYR* | Albinism, oculocutaneous, types IA ; IB | 203100; 606952 | NM_000372.5 | c.1037-7T>A | N/R (intronic) | rs61754381 | LP | 1 | Intronic splice site acceptor | moderate |
| *TYR* | Albinism, oculocutaneous, types IA ; IB | 203100; 606952 | NM_000372.5 | c.1064C>T | p.(Ala355Val) | rs151206295 | PAT | 1 | Missense | moderate |
|  |  |  |  | c.1217C>T | p.(Pro406Leu) | rs104894313 | LP |  | Missense |  |
| *TYR* | Albinism, oculocutaneous, types IA ; IB | 203100; 606952 | NM_000372.5 | c.649C>T | p.(Arg217Trp) | rs63159160 | PAT | 1 | Missense | moderate |
|  |  |  |  | c.1217C>T | p.(Pro406Leu) | rs104894313 | LP |  | Missense |  |
| *MEFV* | Familial Mediterranean fever, AR | 249100 | NM_000243.3 | c.2177T>C | p.(Val726Ala) | rs28940579 | LP | 20 | Missense | mild |

**Table 1. Reported Shared AR variants.**
| Gene | OMIM name | OMIM # | Transcript | HGVSc | HGVSp | dbSNP | ACMG classification | # Affected couples | Effect | Severity Classification\ |
| --- | --- | --- | --- | --- | --- | --- | --- | --- | --- | --- |
| *MEFV* | Familial Mediterranean fever, AR | 249100 | NM_000243.3 | c.2080A>G | p.(Met694Val) | rs61752717 | PAT | 2 | Missense | mild |
| *MEFV* | Familial Mediterranean fever, AR | 249100 | NM_000243.3 | c.2177T>C | p.(Val726Ala) | rs28940579 | LP | 2 | Missense | mild |
|  |  |  |  | c.2080A>G | p.(Met694Val) | rs61752717 | PAT |  | Missense |  |
| *MEFV* | Familial Mediterranean fever, AR | 249100 | NM_000243.3 | c.2040G>C | p.(Met680Ile) | rs28940580 | PAT | 1 | Missense | mild |
|  |  |  |  | c.2177T>C | p.(Val726Ala) | rs28940579 | LP |  | Missense |  |
| *MEFV* | Familial Mediterranean fever, AR | 249100 | NM_000243.3 | c.2040G>C | p.(Met680Ile) | rs28940580 | PAT | 1 | Missense | mild |
|  |  |  |  | c.2080A>G | p.(Met694Val) | rs61752717 | PAT |  | Missense |  |
| *CYP21A2* | Congenital Adrenal hyperplasia | 201910 | NM_000500.9 | c.844G>T | p.(Val282Leu) | rs6471 | PAT | 1 | Missense | mild |
|  |  |  |  | c.1447C>T | p.(Pro483Ser) | rs776989258 | LP |  | Missense |  |
| *CYP21A2* | Congenital Adrenal hyperplasia | 201910 | NM_000500.9 | c.844G>T | p.(Val282Leu) | rs6471 | PAT | 4 | Missense | mild |
| *GNRHR* | Hypogonadotropic hypogonadism 7 without anosmia | 146110 | NM_000406.3 | c.317A>G | p.(Gln106Arg) | rs104893836 | LP | 1 | Missense | mild |

These included *COL6A2* (Ullrich congenital muscular dystrophy 1, n=1), *DOCK7* (developmental and epileptic encephalopathy 23, n=1), *ATP7B* (Wilson disease n=1), *ALDOB* (hereditary fructose intolerance (n=1), *PAH* (phenylketonuria, n=4), and *TGDS* (Catel-Manzke syndrome n=1. In addition, 16.3% (n=8) of couples carried variants associated with moderate disease severity, including *GBA* (Gaucher disease, n=1, (13)), *TYR* (oculocutaneous albinism, n=3), *SEC23B* (congenital dys-erythropoietic anemia I, n=1), *FYCO1* (cataract 1, n=1), *GJB2, GJB6* (digenic *GJB2/GJB6* deafness, n=1), *CLCNKB, CLCNK1* Bartter syndrome (n=1). The remaining 65.3% (n=32) were associated with mild conditions, including *MEFV* (AR Familial Mediterranean fever, n=26), *CYP21A2* (congenital adrenal hyperplasia, n=5), and *GNRHR* (hypogonadotropic hypogonadism without anosmia 7, n=1). In 71.4% (n=35) of the couples, the shared variant was identical, whereas in the remaining 28.6% (n=14) of couples the shared variants differed. None of the couples shared an LP/P CNV or had more than one reported finding.

### Maternal Variants

Out of the 787 female participants, 1.27% (n=10) received a positive maternal result. Among these, 60% (n=6) carried XL variants, 30% (n=3) carried mtDNA variants and one had an AD variant potentially affecting male descendants (Table 2, Figure 1B-C). Three of these findings were CNVs, and none of the identified variants was detected in more than one proband. The XL findings included the following genes: *STS* (X-linked ichthyosis n=2), *ABCD1* (Adrenoleukodystrophy n=1), *CACNA1F* (Aland Island eye disease n=1), *F8* (Hemophilia A n=1), and *FGD1* (Aarskog-Scott syndrome n=1).

**Table 2.** Pathogenic and likely pathogenic maternal-associated variants. List of all maternal findings among female participants with a positive result (n = 10/787; 1.27%), including X-linked (XL), mitochondrial DNA (mtDNA), and autosomal dominant (AD) variants. For each variant, the table reports the gene, number of affected individuals, inheritance pattern, OMIM disease name and number, genomic location or transcript, HGVS cDNA and protein nomenclature (where applicable), dbSNP identifier, ACMG classification, disease severity classification (according to Lazarin et al.(10)), and predicted effect. Abbreviation: ACMG -American College of Medical Genetics.

| SNV |  |  |  |  |  |  |  |  |  |  |  |
| --- | --- | --- | --- | --- | --- | --- | --- | --- | --- | --- | --- |
| Gene | # Affected Individuals | Inheritance | OMIM name | OMIM # | Transcript | HGVSc | HGVSp | dbSNP | ACMG classification | Severity classification | Effect |
| *ABCD1* | 1 | XLR | Adrenoleukodystrophy | 300100 | NM_000033.4 | c.1597A>C | p.(Lys533Gln) | rs781862879 | PAT | Profound | Stop gain |
| *CACNA1F* | 1 | XL, XLR | Aland Island eye disease | 300600 | NM_001256789.3 | c.1138_1141del | p.(Glu380Lysfs*52) | N/A | LP | moderate | Frameshift |
| *F8* | 1 | XLR | Hemophilia A | 306700 | NM_000132.4 | c.1648C>T | p.(Arg550Cys) | rs137852417 | PAT | moderate | Missense |
| *FGD1* | 1 | XLR | Aarskog-Scott syndrome | 305400 | NM_004463.3 | c.1788_1789insGT | p.(Ile597Valfs*46) | N/A | LP | moderate | Frameshift |
| CNV |  |  |  |  |  |  |  |  |  |  |  |
| Gene | # Individuals | Inheritance | OMIM name | OMIM # | Location | ACMG classification | Severity classification | Effect |  |  |  |
| *STS* | 1 | XL | X-linked Ichthyosis | 308100 | chrX:6968336-8138390 | PAT | moderate | Deletion (CNV) |  |  |  |
| *STS* | 1 | XL | X-linked Ichthyosis | 308100 | chrX:6968336-7268302 | PAT | moderate | Deletion (CNV) |  |  |  |
| *DMRT1* | 1 | AD | DOUBLESEX- AND MAB3-RELATED TRANSCRIPTION FACTOR 1; DMRT1 | 602424 (Gene) | chr9:116799-2077776 | PAT | moderate | Deletion (CNV) |  |  |  |
| mtDNA |  |  |  |  |  |  |  |  |  |  |  |
| Gene | # Individuals | Inheritance | OMIM name | OMIM # | Transcript | HGVSc | HGVSp | Location | dbSNP | ACMG classification | Severity classification |
| *MT-RNR1* (100% heteroplasmy) | 1 | mtDNA | COMPLEX I, SUBUNIT ND1; MTND1 | 561000 (Gene) | ENST00000389680.2 | m.1555A>G | N/A | chrMT-1555 A>G | rs2676066 17 | PAT | severe |
| *MT-TS1* (19.56% heteroplasmy) | 1 | mtDNA | TRANSFER RNA, MITOCHONDRIAL, SERINE, 1; MTTS1 | 590080 (Gene) | ENST00000387416.2 | m.7471dup | N/A | chrMT-7465 A>AC | rs1110333 19 | PAT | severe |
| *MT-ND4* (100% heteroplasmy) | 1 | mtDNA | COMPLEX I, SUBUNIT ND4; MTND4 | 516003 (Gene) | ENST00000361381.2 | m.11778G>A | p.(Arg340His) | chrM-11778 G>A | rs1994761 12 | PAT | severe |

The mitochondrial findings involved the following variants: m.1555A>G (n=1) with 100% heteroplasmy, associated with hearing loss following exposure to aminoglycoside antibiotics during childhood. It has been predominantly reported in homoplasmic individuals (14–16); m.7471dup (n=1) with 19.56% heteroplasmy, most commonly associated with sensorineural hearing loss however affected individuals can also have other features, including MELAS, cerebellar ataxia, myoclonus, epilepsy, white matter disease, intellectual disability, and more; and m.11778 G>A (n=1) with 100% heteroplasmy, associated with Leber hereditary optic neuropathy, among other conditions.

Additionally, one AD variant was identified as a 2Mb deletion on chromosome 9, encompassing the entire *DMRT1* gene. Deletions in the 9p24.3 region have been reported in individuals with disorders of sexual development, presenting a spectrum ranging from 46,XY chromosomal males with female phenotypes to males with azoospermia (17,18).

While excluded from the primary maternal results, the common *G6PD* variant (NM_001360016.2:563C>T, p.(Ser188Phe) was identified in 6.1% (96/1574) of the MJC cohort (66 heterozygous and 2 homozygous females, and 28 males). Given that hemizygous males who inherit this variant have an increased risk of G6PD deficiency, it was reported to female carriers as an additional maternal finding but was not included in the primary set of maternal results.

### Exome Diagnostic Yield vs. Other Screening Panels

Overall, the ES identified 59 at-risk couples: 49 couples with shared AR variants and 10 couples with maternal-linked variants, across 23 nuclear genes and three mitochondrial genes. For a direct comparison with standard carrier screening panels, which assess AR conditions only, the analysis was restricted to the 49 couples carrying shared AR variants. When comparing these findings to the three standard carrier screening modalities -ACMG Tier 3, the Pan-ethnic Panel, and the Dor Yeshorim extended panels, ES identified AR variants in nine nuclear genes that would have otherwise gone undetected. In total, ES revealed 12 additional at-risk couples with AR conditions, including those affected by severe conditions such as Catel-Manzke syndrome and DEE 23, caused by the *TGDS* and *DOCK7* genes, respectively.

Specifically, the ACMG Tier 3 panel and the Pan-ethnic panel would have failed to identify 10 and 15 at-risk couples with AR conditions, respectively. The Dor Yeshorim panel would have detected only a single couple with the *COL6A2* c.1402C>T variant, thereby missing 48 couples with AR variants, 13 of whom carried variants associated with moderate to profound conditions. Table 3 summarizes this comparison and, for demonstration purposes, highlights only AR variants classified as moderate to profound.

**Table 3.** Exome sequencing diagnostic yield compared with standard carrier screening panels (autosomal recessive conditions). Comparison of at-risk couples identified by ES versus three routinely used carrier-screening modalities (ACMG Tier 3 panel, Pan-ethnic panel, and Dor Yeshorim published panel from 2021). To enable a direct like-for-like comparison, this table is restricted to autosomal recessive (AR) conditions; X-linked and mitochondrial variants are excluded as they fall outside the scope of standard carrier screening panels. The table lists for each gene the variant combinations, mode of inheritance, severity classification, and the number of at-risk couples detected by ES. Only variants associated with moderate to profound conditions are included for demonstration purposes. Abbreviations: ACMG - American College of Medical Genetics.

| Gene | Variant | ACMG Tier 3 Panel | Pan Ethnic Panel | Dor Yeshorim Panel | Inheritance | Severity classification | Couples (C) / Individuals (I) |
| --- | --- | --- | --- | --- | --- | --- | --- |
| *ALDOB* | c.448G>C | Yes | Yes | No | AR | severe | 1 C |
| *ATP7B* | c.845del | Yes | Yes | No | AR | severe | 1 C |
| *CLCN1* | c.378dup | Yes | No | No | AR | moderate | 1 C |
|  | c.501C>G |  |  |  |  |  |  |
| *CLCNK1* | exons 4-20 deletion | *No* | *No* | No | AR | moderate | 1 C |
| *CLCNKB* | exons 1-3 deletion | *No* | *No* | No |  |  |  |
|  | c.1830G>A |  |  |  |  |  |  |
| *COL6A2* | c.1402C>T | *No* | *No* | Yes | AR | severe | 1 C |
| *DOCK7* | c.1832_1835del | *No* | *No* | No | AR | profound | 1 C |
| *FYCO1* | c.3150+1G>T | *No* | *No* | No | AR | moderate | 1 C |
| *GBA* | c.1448T>C | Yes | Yes | No | AR | moderate | 1 C |
| *GJB2* | c.101T>C | Yes | Yes | No | AR | moderate | 1 I |
| *GJB6* | Exons 1-5 deletion | *No* | *No* | No | AR | moderate | 1 I |
| PAH | c.1066-11G>A | Yes | Yes | No | AR | severe | 4 C |
|  | c.898G>T |  |  |  |  |  |  |
|  | c.688G>A |  |  |  |  |  |  |
|  | c.727C>T |  |  |  |  |  |  |
|  | c.1208C>T |  |  |  |  |  |  |
|  | c.829T>C |  |  |  |  |  |  |
|  | c.1243G>A |  |  |  |  |  |  |
| SEC23B | c.325G>A | No | No | No | AR | moderate | 1 C |
| TGDS | c.281_282del | No | No | No | AR | severe | 1 C |
| TYR | c.1037-7T>A | Yes | No | No | AR | moderate | 3 C |
|  | c.1064C>T |  |  |  |  |  |  |
|  | c.1217C>T |  |  |  |  |  |  |
|  | c.649C>T |  |  |  |  |  |  |

### Secondary Findings

A total of 21% of individuals opted out of receiving secondary findings. To ensure accurate calculation of secondary finding rates, all tested individuals were included in the analysis. A total of 2.85% (n=45) individuals were found to carry 29 variants across 14 different genes included in the ACMG SF v3.3 list (11). Among the secondary findings, 53.33% (n = 24) of the variants were associated with cardiac disorders, encompassing six genes: *LDLR* (familial hypercholesterolemia, n = 8), *TTN* (dilated cardiomyopathy, n = 5), *MYH7* (hypertrophic cardiomyopathy, n = 4), *DSP* (arrhythmogenic right ventricular cardiomyopathy n = 4), *KCNQ1* (long QT syndrome, n = 1), *KCNH2* (long QT syndrome, n = 1) and *TGFBR1* (Loeys–Dietz syndrome type 1, n=1) (Table 4, figure 1D).

**Table 4.**
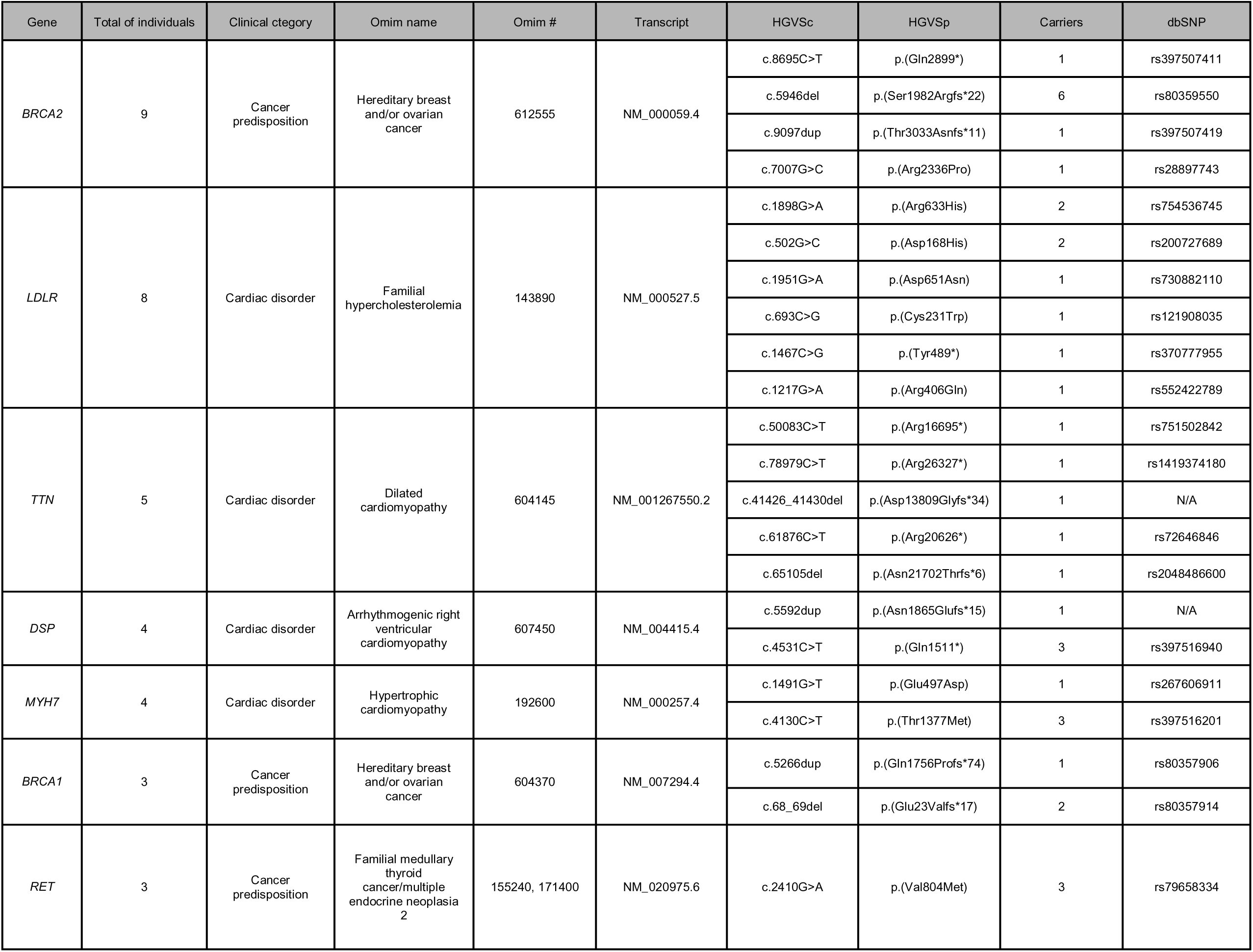
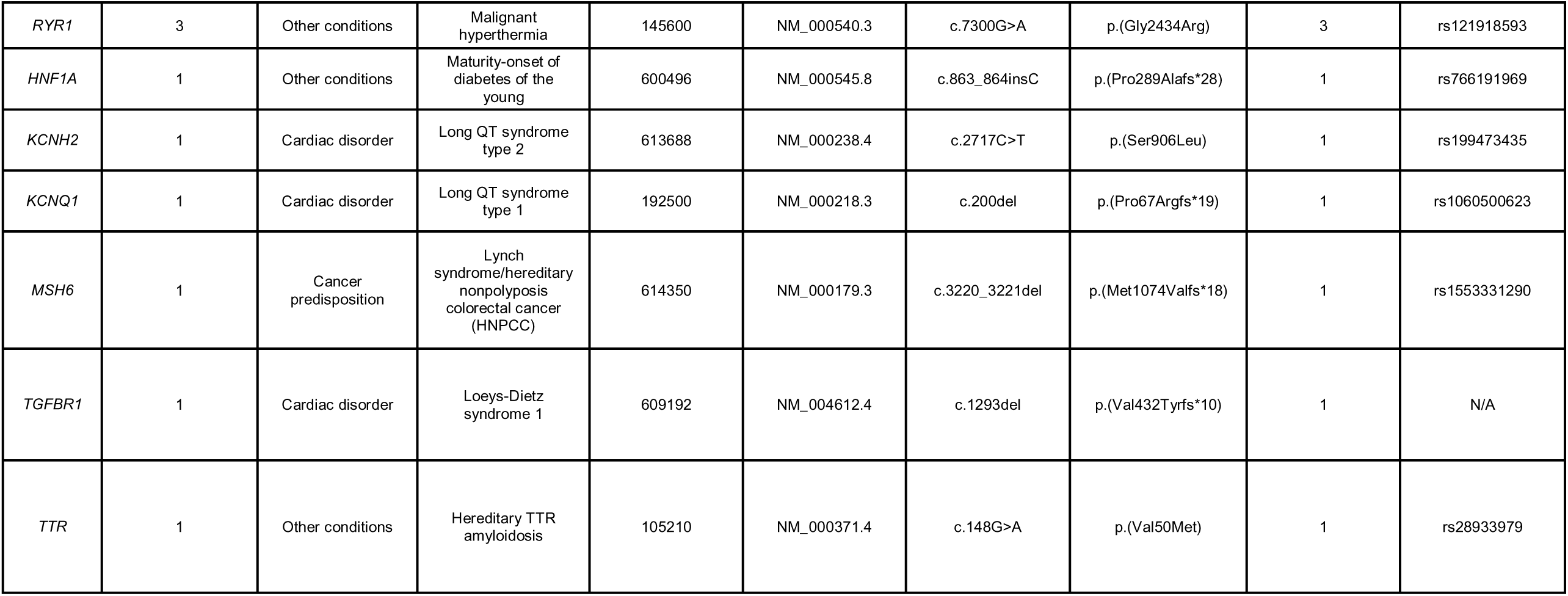
ACMG secondary findings. Summary of all pathogenic and likely pathogenic variants reported under the ACMG SF v3.3 actionable gene recommendations, identified in 45 individuals (2.85% of the cohort). The table lists, for each gene, the associated condition, total number of carriers, reference transcript, HGVS cDNA and protein nomenclature, dbSNP identifier (where available), and disease severity classification (according to Lazarin et al.(10)), grouped into cardiac disorders, cancer-predisposition syndromes, and other actionable conditions.

Variants associated with cancer predisposition genes accounted for 35.5% (n = 16) of the SF and were identified in four genes: *BRCA2* (hereditary breast and ovarian cancer, n=9), *BRCA1* (hereditary breast and/or ovarian cancer, n=3), *RET* (multiple endocrine neoplasia types IIA/IIB, n=3), and *MSH6* (Lynch syndrome, n=1).

The remaining 11.11% (n=5) corresponded to variants associated with other conditions, including *RYR1* (malignant hyperthermia, n=3), *HNF1A* (Maturity-onset of diabetes of the young n=1), and *TTR* (hereditary TTR amyloidosis, n=1) (Table 4, Figure 1D).

Variants in the *BRCA2* gene were the most frequent among all SFs, identified in nine carriers. Six of these individuals carried the *BRCA2* Ashkenazi founder variant (*c.5946delT*), while three variants were novel (Table 4, Figure 1D). Interestingly, three out of the six *BRCA2* Ashkenazi founder variant carriers did not self-identify as Ashkenazi, but as Syrian.

### Genomic Profiling of the Mexican Jewish Community

To evaluate the necessity of ES for PCS in the MJC, we constructed and analyzed an individual-level database of heterozygous LP/P variants. Briefly, the database included: (i) variants with >20 homozygous occurrences in gnomAD v2.1.1 that are classified as P/LP in public databases such as ClinVar or the Israeli National Genetic Database (INGD); and (ii) rare LOF variants in AR and XL genes, defined as those with ≤2 homozygous and ≤700 heterozygous occurrences in gnomAD V2.1.1.

### Rare LP/P Variants in the Population

To assess the genetic distinctiveness of the MJC, we extracted LP/P variants with an allele frequency ≤0.005 (∼1/200) in gnomAD v2.1.1. A total of 920 such variants were identified, of which 146 were novel and entirely absent from gnomAD. Among these novel variants, 80 were observed in a single individual, while the remainder were detected in 2–27 individuals.

Of the 920 rare LP/P variants, six were LOF variants that have not been previously reported in the literature or submitted to ClinVar (Supplementary Table 1).

### Frequent LP/P variants among the population

The 20 most frequent variants identified in the MJC and their corresponding global frequencies are detailed in Supplemental Table 2. These high-frequency alleles were involved in 69.4% (n=34) of the 49 shared carrier cases identified in this study.

The *MEFV* gene exhibited the highest prevalence, with variants identified in 217 individuals (13.7% of the cohort). While shared *MEFV* variants were detected in 3.3% of the couples (26/787, Table 1), one of the partners had a biallelic *MEFV* variant. ES also identified nine additional individuals with biallelic *MEFV* variants (five homozygous, four compound heterozygous). Although these individuals self-reported as healthy, they require further clinical examination to establish a definitive diagnosis-a finding that aligns with the significant carrier frequency of Familial Mediterranean Fever in this population.

Other prevalent variants in the cohort included *GNRHR* c.317A>G (3.3%; 52/1574), *COL6A2*c.1402C>T (1.7%; 27/1574), and *CYP21A2* c.844G>T (0.3%; 5/1574). Shared status for these genes, along with *TYR* variants, accounted for most of the remaining high-frequency positive findings in the cohort.

## DISCUSSION

This study demonstrates the significant utility of ES for PCS within the Mexican Jewish Community. Shared carrier screening identified 49 at-risk couples for AR conditions, with 34.6% (17/49) of these cases involving moderate to profound disorders. Beyond shared AR status, the inclusion of maternal-linked findings -spanning X-linked, mitochondrial, and specific AD variants -identified an additional 10 women at risk. When factoring in SFs, nearly 10% (153/1574) of the total cohort received a clinically actionable result, underscoring the broad impact of this screening approach.

The genetic landscape of the MJC is uniquely shaped by its history. While the global Jewish population shares ancient Levantine roots, the MJC has been forged through distinct migratory waves of Ashkenazi, Sephardi, and Syrian-Mizrahi groups (19,20). The relative genetic isolation and endogamous practices within these sub-communities have created a heterogeneous genetic architecture characterized by specific founder effects and genetic drift. Our findings highlight this complexity; for example, the identification of the Ashkenazi *BRCA2* founder variant in individuals who did not self-identify as Ashkenazi suggests either deeper historical admixture between Jewish sub-groups or that these variants predate the diasporic split. This finding is further supported by the Dor Yeshorim study, which found relatively high Syrian Jewish carrier frequencies (>0.68%) for well-established Ashkenazi Jewish variants (12).

A primary strength of this study is the evidence it provides for the superior diagnostic yield of ES over conventional panels. Our analysis revealed that over 40% of the positive cases would have been missed by ACMG Tier 3 guidelines, and nearly 60% would have gone undetected by standard pan-ethnic panels. The Dor Yeshorim panel reported previously was found to detect a single couple. This reflects the nature of the program, as variants are added to the tested panel only when they meet strict criteria for high carrier frequency, severe early-onset clinical impact, and the availability of reliable, well-validated assays for accurate detection. Of note, the compared panel is outdated, and the number of variants has since been expanded to increase the proportion of detectable at-risk couples.

Notably, ES identified severe conditions, such as Catel-Manzke syndrome and DEE 23, that are not captured by routine community screening programs. This discrepancy emphasizes that guidelines based predominantly on U.S. and European data may fail to account for the population-specific pathogenic variants found in genetically diverse or isolated groups like the MJC. Accordingly, several large-scale studies have demonstrated that the spectrum and frequency of pathogenic variants differ significantly across populations. For instance, *Tong et al.* (2022) and *Chau et al.* (2022) conducted expanded carrier screening using medical ES in over 4,000 and 1,500 individuals from China, respectively, reporting carrier rates exceeding 90% and substantial differences in gene distribution compared with Western cohorts. Similarly, *Belnap et al.* (2025) applied exome and genome sequencing for carrier screening in the United States, highlighting variability even within subpopulations. These findings emphasize the need to generate population-specific data in regions such as Latin America, where both genetic admixture and founder effects play an important role in disease prevalence and variant interpretation (8,21–23).

Furthermore, the simultaneous detection of SFs transforms PCS from a purely reproductive tool into a proactive instrument for precision preventive medicine. By identifying variants associated with hereditary cancers and cardiac conditions in healthy young adults, ES provides an opportunity for early clinical intervention and family-wide risk assessment. This is particularly relevant in the MJC, where we identified a 1.01% carrier frequency for cancer-predisposition genes, including both known founder variants and novel variants.

Despite these advantages, the implementation of ES in underrepresented populations involves several critical challenges. The identification of variants of uncertain significance (VUS) is more frequent when comparative population data is limited, which can lead to clinical ambiguity and patient anxiety. While ES is robust for detecting SNVs, the potential for false-positive results in CNVs detection remains a limitation that often requires secondary confirmation through additional methodologies.

Furthermore, although ES is intended to capture the entire exome, it typically misses approximately 6% of coding regions and is generally unable to detect variants located in intronic or regulatory sequences. ES also possesses significant "blind spots" for specific classes of genetic disorders, including trinucleotide repeat expansions (such as Fragile X syndrome) and genes with complex genomic architectures or highly similar pseudogenes, such as *SMN1* (spinal muscular atrophy) or specific regions of *CYP21A2* and *GBA*. Because these variants are technically challenging to resolve using standard ES pipelines, they often necessitate specialized ancillary testing, such as MLPA or PCR-based assays. As a result, while a negative ES result substantially lowers reproductive risk, it does not completely eliminate the risk for the entire spectrum of conditions usually covered in expanded carrier screening. Future transitions to whole-genome sequencing (WGS) could eventually address these gaps by providing more reliable CNV analysis and better coverage of regulatory regions.

Finally, while the study is limited by its non-randomized, voluntary enrollment and the lack of long-term follow-up on reproductive outcomes, the large cohort of healthy individuals provides a powerful cross-section of the MJC’s genetic risk distribution. These results not only support the implementation of exome-based PCS in the MJC but also serve as a model for other endogamous or understudied populations globally.

## Data Availability

All analyzed data contain patients' personal data and are stored in accordance with institutional regulations. Anonymized data are available upon request.

## AUTHOR APPROVAL

All authors have approved the manuscript and its submission.

## ACKNOWLEDGEMENTS

We acknowledge the valuable contribution of the Jewish Community in Mexico Zug Project, the volunteers who have made this project possible: Cessy Metta, Gabriela Costi, Susy Maya, Alberto Kichik, Simon Credi, Victor Achar, Pepe Cohen, Elías Zaga, Alan Abadi, Ariela Jinich, Andrea Hirschhorn, Keren Goldrin, Elvira Edid, Ariela Souroujon, Irene Dichi-Javnozon, Adela Kanan-Masri, and Sociedad de Beneficencia Alianza Monte Sinaí.

We would also like to thank the rabbis of the Puah Institute in Mexico, Rabbi David Jabbaz and Rabbi Daniel Credi, for their significant contributions to raising awareness of genetics within the community, and especially to all the consented couples.

## ETHICS APPROVAL STATEMENT

The study was conducted in accordance with IRB-approved protocol 0306-10-HMO given by Hadassah Medical Center.

