## Supplemental Tables for "Exome-based Preconception Carrier Screening for the Mexican Jewish Community; Yields and Implications"

Supplemental table 1

| Gene | Omim name | Omim # | Transcript | HGVSc | HGVSp | dbSNP | ACMG Classification | Zygosity | # of carriers | GnomAD v2.1.1 HOM | GnomAD v2.1.1 HET | Effect | Severity Classification\ |
| --- | --- | --- | --- | --- | --- | --- | --- | --- | --- | --- | --- | --- | --- |
| *TGDS* | Catel-Manzke syndrome; AR | 616145 | NM_014305.4 | c.281_282delTA | p.(Ile94fs*19) | rs1889177568 | LP | HET | 8 | N/A | N/A | Frameshift | severe |
| *ALKBH8* | Intellectual developmental disorder 71; AR | 618504 | NM_138775.3 | c.1657_1658delTCinsATG | p.(Ser553fs) | N/A | LP | HET | 5 | N/A | N/A | Frameshift; non synonimous | severe |
| *CUBN* | Chronic benign Proteinuria; AR, Imerslund-Grasbeck syndrome 1; AR | 618884; 261100 | NM_001081.4 | c.8911delT | p.(Ser2971fs*3) | N/A | LP | HET | 4 | N/A | N/A | Frameshift | moderate |
| *DNAH9* | Primary ciliary dyskinesia; AR | 618300 | NM_001372.4 | c.4844_4847delTCCT | p.(Leu1616fs*27) | rs746250627 | LP | HET | 3 | 0 | 3 | Frameshift | moderate |
| *NCAPG2* | Khan-Khan-Katsanis syndrome; AR | 618460 | NM_017760.7 | c.2698_2699insTAGCCATCT | p.(Gln900delinsLeuAlaIleTer) | N/A | LP | HET | 1 | N/A | N/A | Codon insertion; Stop gain | severe |
| *ASPM* | Primary microcephaly 5; AR | 608716 | NM_018136.5 | c.9165_9166insCTATAATGT | p.(Ala3055_Ala3056insLeuTerCys) | N/A | LP | HET | 1 | N/A | N/A | Codon insertion; Stop gain | severe |

**Supplementary Table 1. Novel rare loss‑of‑function variants identified in the Mexican Jewish community database.**

List of seven heterozygous rare LP/P loss‑of‑function variants that are absent from public variant repositories. For each variant, the table provides gene symbol, OMIM disease name and number, reference transcript, HGVS cDNA and protein nomenclature, zygosity, number of carriers in the cohort, gnomAD homozygous and heterozygous counts (v2.1.1), predicted functional effect (e.g., frameshift, stop‑gain), and assigned disease‑severity classification.

Supplemental table 2

| # | Gene | Omim name | Omim # | Transcript | HGVSc | HGVSp | dbSNP | ACMG classification | Effect | alt allele in MJC | Global AF (GnomAD v.2.1) | Ashkenazi AF (GnomAD v2.1) | Severity classification |
| --- | --- | --- | --- | --- | --- | --- | --- | --- | --- | --- | --- | --- | --- |
| 1 | *MEFV* | Familial Mediterranean fever; AR | 249100 | NM_000243.3 | c.2177T>C | p.(Val726Ala) | rs28940579 | LP | missense | 0.054955527 | 0.002167119 | 0.039789641 | mild |
| 2 | *G6PD* | Glucose‑6‑phosphate dehydrogenase deficiency; XR, Chronic nonspherocytic hemolytic anemia; XR, Favism; XR | 300908, 305900, 134700 | NM_001360016.2 | c.563C>T | p.(Ser188Phe) | rs5030868 | LP | missense | 0.040660737 | 0.002553791 | 0.006014435 | moderate |
| 3 | *CYP21A2* | Adrenal hyperplasia, congenital, classic 21‑hydroxylase deficiency; AR, Adrenal hyperplasia, congenital, nonclassic; AR | 201910, 202010 | NM_000500.9 | c.844G>T | p.(Val282Leu) | rs6471 | PAT | missense | 0.028589581 | 0.00528419 | 0.023287362 | mild |
| 4 | *GNRHR* | Hypogonadotropic hypogonadism 7; AR | 146110 | NM_000406.3 | c.317A>G | p.(Gln106Arg) | rs104893836 | LP | missense | 0.016518424 | 0.002841949 | 0.005857824 | mild |
| 5 | *PMM2* | Congenital disorder of glycosylation type Ia; AR | 212065 | NM_000303.3 | c.422G>A | p.(Arg141His) | rs28936415 | PAT | missense | 0.01111817 | 0.004067906 | 0.00770416 | severe |
| 6 | *SLC10A1* | Hypercholanemia; AR, Rotor syndrome; AR | 607748, 237450 | NM_003049.4 | c.755G>A | p.(Arg252His) | rs147226818 | LP | missense | 0.009212198 | 0.00026168 | 0.000436967 | moderate |
| 7 | *ERCC2* | Xeroderma pigmentosum group D; AR, Trichothiodystrophy 1; AR, Cerebro‑oculo‑facial‑skeletal syndrome 2; AR | 278730, 601675, 610756 | NM_000400.4 | c.466C>T | p.(Arg156*) | rs151235136 | PAT | stop gain | 0.008894536 | 1.19737E-05 | 0 | severe |
| 8 | *COL6A2* | Bethlem myopathy 1; AD/AR, Ullrich congenital muscular dystrophy 1; AR, Myosclerosis myopathy; AR | 158810, 254090, 255600 | NM_001849.4 | c.1402C>T | p.(Arg468*) | rs374669775 | PAT | stop gain | 0.008576874 | 1.21063E-05 | 0.0001001 | severe |
| 9 | *STAR* | Adrenal hyperplasia, congenital, lipoid; AR, Adrenal hyperplasia, congenital, nonclassic lipoid; AR | 201710, 614080 | NM_000349.3 | c.178+1G>C | N/A | rs1554503011 | LP | splice site donor | 0.008576874 | 0.000001239 | 0 | severe |
| 10 | *TYR* | Oculocutaneous albinism type 1A; AR, Oculocutaneous albinism type 1B; AR | 203100, 606952 | NM_000372.5 | c.1037-7T>A | N/A | rs61754381 | LP | Intronic splice site acceptor | 0.008259212 | 0.000949717 | 0.015239044 | moderate |
| 11 | *G6PC1* | Glycogen storage disease Ia; AR | 232200 | NM_000151.4 | c.247C>T | p.(Arg83Cys) | rs1801175 | PAT | missense | 0.007623888 | 0.000572674 | 0.006646825 | severe |
| 12 | *MYO6* | Deafness, autosomal dominant 22; AD, Deafness, autosomal recessive 37; AR | 600970, 607821 | NM_004999.4 | c.2742_2743insA | p.(Gln918fs*24) | rs551348450 | P | frameshift | 0.007306226 | 0.0003225 | 0.0003484 | moderate |
| 13 | *GJB2* | Deafness, autosomal recessive 1A; AR, Deafness, autosomal dominant 3A; AD, Keratitis‑ichthyosis‑deafness syndrome; AD | 220290, 601544, 148210 | NM_004004.6 | c.35delG | p.(Gly12fs*2) | rs80338939 | P | frameshift | 0.006988564 | 0.006188 | 0.003378 | moderate |
| 14 | *GAA* | Glycogen storage disease II (Pompe disease); AR | 232300 | NM_000152.5 | c.-32-13T>G | N/A | rs386834236 | P | intronic | 0.006670902 | 0.003401 | 0.005541 | severe |
| 15 | *ZNF469* | Brittle cornea syndrome 1; AR | 229200 | NM_001367624.2 | c.19C>T | p.(Arg7*) | rs1004428835 | LP | stop gain | 0.006670902 | 0.0003071 | 0 | mild |
| 16 | *FAM161A* | Retinitis pigmentosa 28; AR | 606068 | NM_001201543.2 | c.1567C>T | p.(Arg523*) | rs202193201 | PAT | stop gain | 0.006670902 | 0.00004629 | 0 | moderate |
| 17 | *CYP21A2* | Adrenal hyperplasia, congenital, classic 21‑hydroxylase deficiency; AR, Adrenal hyperplasia, congenital, nonclassic; AR | 201910, 202010 | NM_000500.9 | c.1447C>T | p.(Pro483Ser) | rs776989258 | P | missense | 0.00635324 | 0.0005244 | 0.000632 | mild |
| 18 | *HESX1* | Septo‑optic dysplasia; AR/AD, Pituitary hormone deficiency, combined 5; AR | 182230, 613038 | NM_003865.3 | c.475C>T | p.(Arg159Trp) | rs770886420 | LP | missense | 0.00635324 | 0.00001993 | 0 | moderate |
| 19 | *WDR35* | Cranioectodermal dysplasia 2 (Sensenbrenner syndrome); AR, Short‑rib thoracic dysplasia 7; AR | 614378, 614091 | NM_020779.4 | c.1694_1695dupTA | p.(Thr566fs*1) | rs766096320 | LP | frameshift | 0.00635324 | 0.0000398 | 0.0009923 | severe |
| *20* | *RSPH1* | Primary ciliary dyskinesia 24; AR | 615874 | [NM_080860.4](http://www.ncbi.nlm.nih.gov/snp/rs151107532) | c.275-2A>C | N/A | [rs151107532](http://www.ncbi.nlm.nih.gov/snp/rs151107532) | PAT | splice site acceptor | 0.00635324 | 0.0003535 | 0 | severe |

**Supplementary Table 2. Most frequent pathogenic and likely pathogenic variants in the Mexican Jewish community cohort.**

List of the 20 LP/P variants with the highest allele frequencies in the study population, including gene symbol, reference transcript, HGVS cDNA and protein nomenclature, dbSNP identifier, ACMG classification, predicted molecular effect, alternate‑allele frequency in the Mexican Jewish cohort, and corresponding global and Ashkenazi Jewish allele frequencies from gnomAD v2.1.
